# The UK Covid-19 lockdown weakened in April and May 2020: implications for the size of the epidemic and for outcomes had lockdown been earlier

**DOI:** 10.1101/2020.07.04.20146340

**Authors:** Anthony Lander

## Abstract

The number of active cases in the UK Covid-19 epidemic, the case fatality rate, the susceptible proportion of the population, and how well the lockdown was maintained during April–May 2020 are unknown. These four have a relationship with the shape of the daily mortality curve once one considers the intervals from infection to death or recovery. Without an understanding of this relationship we cannot say that an earlier lockdown would have saved lives. Using a small stochastic model, the lockdown had to be weakened, in April and May, for simulated deaths to match ongoing actual daily deaths. Google mobility data was found to be consistent with the weakening required in the model with similar changes from baseline in time and magnitude. If in an earlier lockdown, mobility and interactions would have followed a similar course, then with a large epidemic curve an earlier lockdown might be associated with many more deaths than some currently believe. This was confirmed in the stochastic model and in two modified SIR models of epidemics of various sizes. The first SIR model had a fixed period to recovery and the second used random periods, both models had random periods to death. Weakening of the mitigations was required to tune the output in large but not in small epidemics. This gives weight to the epidemic having affected many more individuals than some reports currently suggest. In both one and two-week earlier lockdowns, total deaths were found to depend on the size of the epidemic and to vary from 2,000–49,000 deaths. There was a linear relationship between the peak proportion of the population infected and the reciprocal of the case fatality rate. This work questions the low prevalence of < 0.1%, reported by the Office for National Statistics in May and June 2020, since to accommodate a weakening lockdown, the shape of the daily mortality curve, and an acceptable case fatality rate a much larger epidemic curve is required.

## 1. Introduction

A plot of the time course of the proportion of the population who are currently infected—active cases—is known as the epidemic curve. During mitigated Covid-19 epidemics nations have counted deaths in different ways and the UK have altered its reporting. However, if a mortality data set has been consistently ascertained then the shape of that data maintains some fidelity with its antecedents and inferences may be generalisable. The principal antecedents are those elements that underly the mechanics generating the epidemic curve and the timing and effectiveness of the mitigations and importantly any unintended weakening of the mitigations. In the absence of a tight confidence interval on the case fatality rate the shape, but not the size, of the epidemic curve can be estimated from the mortality data. The size of the infected population may not be required to answer some questions. However, it is required if one asks ‘What might have happened if the mitigations had been introduced earlier?’ This requires an understanding of the size of the epidemic curve since the efficacy of any weakening in the mitigations will be sensitive to the susceptible proportion of the population. Importantly, the size of the epidemic curve and knowing the residual susceptible population are central to understanding the risks of a second-wave and planning relaxation of lockdown rules.

This author previously reported a stochastic model which simulated mainly UK in-hospital deaths [1]. That model was revised to include out-of-hospital deaths after Public Health England (PHE) changed its reporting system in late April 2020 [2]. Subsequent but modest revisions were issued [3]. In this work a first SIR model [A] used a fixed period to recovery of 12.8 days and PHE data up to a revision in late June whilst the second SIR model [B] which used a random period to recovery was tuned following a PHE revision accessed on 26 June [4].

A UK national newspaper, reported that 75% of UK Covid-19 deaths might have been avoided if the lockdown had begun one week earlier [5]. Dagpunar, using a case fatality rate of 1%, describes a SEIR (Susceptible, Exposed, Infected, Removed) model which generated 39,000 deaths for a mitigated epidemic, reducing to around 11,000 if mitigated one week earlier, see Table 1 [6].

**Table 1.** Results from modeling by Dagpunar **[6]**.

|  | Deaths | Peak deaths | Final Susceptible population |
| --- | --- | --- | --- |
| Unmitigated | 634,000 | 18,000 | 4.8% |
| UK-like lockdown | 39,000 | 930 | 94% |
| One-week earlier lockdown | 11,200 | 260 | 98% |

The UK introduced its lockdown when a total of 359 deaths had been reported, whilst Germany locked down after reporting a total of 86 deaths. On 17 March there had been a total of 81 deaths in the UK. It is an important question to ask; ‘What might have happened—if all else remained the same—but the lockdown was announced at a different time?’ This work simulated earlier and later lockdowns starting from 3–20 March 2020 using the previously described stochastic model.

In this stochastic model, just as in a SIR model, the size of the susceptible proportion of the population at any one time influences the efficacy of an intervention, be it a mitigation or a weakening of a lockdown. For this reason, if a set of parameters influencing a model’s behaviour are allowed to act on a population with a substantially different susceptible proportion then their influence will also be quite different. Caution is therefore needed in moving parametric changes earlier or later in an epidemic if the true magnitude of the epidemic curve is not known. Early in an epidemic when exponential growth is seen then the size of the susceptible population may be ignored but this does not apply later on.

Though we know something of the shape of the mortality data, and from this we can infer something of the shape of the epidemic curve, we cannot determine the magnitude of the epidemic curve without other information. The areas under the epidemic curve and the mortality curve give us the case fatality rate.

In this author’s previous simulations around 20% of the population were infected at the peak, but this is higher than the consensus of current opinion. According to the Office for National Statistics (ONS) between 17–30 May 2020, an average of 0.10% of the population in households had Covid-19 (95% CI: 0.05% to 0.18%) [9]. A second estimate between 25 May to 7 June found 0.06% (95% CI : 0.02% to 0.12%) of the population in England had Covid-19 [7]. This second estimate was based on 11 individuals in 8 households from a total of 19,933 participants.

In this work modelling is presented for different sized epidemics. The early parameters are the same, as the susceptible populations have not fallen sufficiently, but later parameters necessarily differ.

### A thought experiment

Though it is unlikely that the UK epidemic was initiated with a single index case we will consider this in a simple thought experiment. If the rising phase is neither mitigated nor yet influenced by the falling susceptible population then it is exponential, and if the doubling was every 3 days then there would have been 25.5 days to get to 360 deaths at lockdown on 23 March placing the first death on 27 February. PHE record the first UK death on 5 March. The antecedent infections (perhaps 20 days before the 23 March) would, at 3 March, have been a magnitude larger related to the case fatality rate (CFR). If we imagine CFR’s of 1:1000 and 1:100 these place the hypothetical first cases on the 7 and 18 January respectively, see Figure 1.

**Figure 1.**
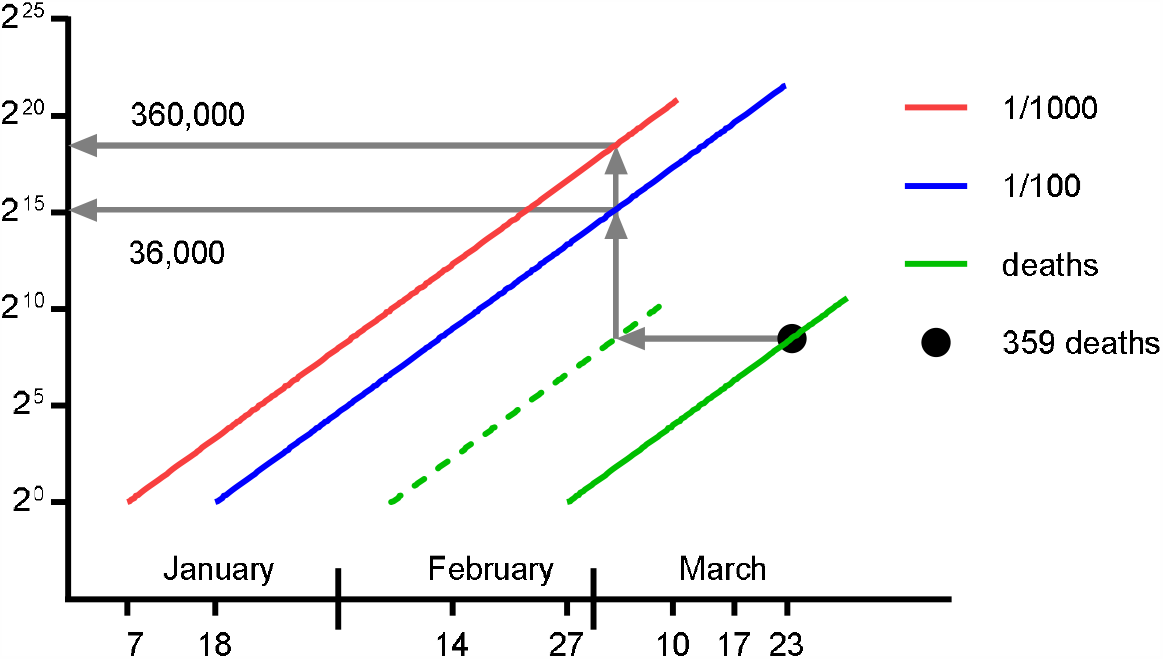
A thought experiment is illustrated. Assuming a 3-day doubling in deaths (green) arising from antecedent infections 20 days previously (dashed green line) producing a total of 359 deaths on 23 March the parent populations can be imagined for different case fatality rates. This crude approximation places the start of the epidemic curves in early January.

## 2. Materials and Methods

Two modified SIR epidemics [A] and [B] were given start dates from 2–23 January. In [A] there was a fixed period of 12.8 days to recovery for those becoming immune and in [B] a randomly chosen period from two lognormal distributions. In both [A] and [B] the period to mortality was randomly chosen. Each epidemic had its weakening tuned such that the daily mortality represented actual daily mortality with social distancing and lockdown taking place on 17 and 24 March. Tuning was a visual fit with one run of the model. Once tuned, each epidemic was re-run with social distancing taking place either one or two-weeks week earlier (10 or 3 March). Lockdown likewise was earlier on the 17 or 10 March.

### The stochastic model

The stochastic model has been previously described [1,2]. The model is written in Microsoft Excel VBA v7·1 operating in Windows 10. Individuals were assigned an age, sex, risk of mortality if infected, a measure of daily viral exposure, a susceptibility factor, an incubation period, and two contagious periods. One contagious period was for mild illness and the other for serious, critical or fatal illness. Each individual had a 2-dimensional location, a status for symptoms, immunity and a flag for alive/dead. Individuals had two clocks; one for an assigned contagious period and one for a virtual twin, 80% of whom were assigned a lognormal fatal illness contagious period to represent in-hospital deaths (meanlog 2.99, sdlog 0.223), and 20% a shorter, lognormally distributed period to represent out-of-hospital deaths (meanlog 2.639, sdlog 0.2). The 80:20 ratio matches the PHE ascertainment of deaths [2]. Incubation periods were based on Covid-19 data (meanlog 1.621, sdlog 0.418) [8]. Age and sex followed the 2011 UK census. Knowing the age and sex, an individual was designated to die, if infected, based on a probability taken from the Chinese Centre for Disease Control and Prevention [9]. The demographic details do not influence the outputs reported in this paper but were maintained in case they prove useful in later developments.

A random 5% were assigned to be critically ill, and together with the dying individuals, this group had the longer contagious period (meanlog of 2.99, sdlog of 0.223). A random, normally distributed (mean 1, sd 0.2) susceptibility factor *s* was chosen. Randomly located and randomly moving individuals become infected if they exceed a daily infectious dose based on their cumulative proximity to contagious near neighbours and their susceptibility factor. The exposure, consequent upon an interaction between a susceptible individual *i* and a contagious individual *j*, had a relationship with the inverse square distance separating the individuals *d*_*i,j*_. All separations *d*_*i,j*_ less than 0·2 metres were assumed to be at 0·2 metres. Separations exceeding 5 m were ignored. When social distancing was applied, separations less than 2 m were treated as if at 2 m, this rule was applied for a percentage *x* of interactions designated *SD*_*x*_. If for any susceptible *i*, on any day, 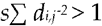 then the individual acquired an infection. One day elapsed between infection and symptoms. A percentage *x*, of symptomatic individuals were confined, making no movements, and designated *C*_*x*_, representing isolation. Mobile individuals could still come close to the confined. Individuals moved *n* times a day designated *M*_*n*_, in a random walk (step-size 10 times the output of an inverse cumulative normal distribution with mean 0 m, sd 5 m). Individuals returned to their origins at the end of each day. Mobile individuals meeting boundaries were reflected to stay within the boundaries. In these simulations 2000 individuals were placed randomly in a 490 × 490 m square. Only small populations were needed since virtual twins were followed to death.

The model was parameterised as follows; *M*_*n*_ with *n* a surrogate for movements, a larger *n* simply representing more movements, *SD*_*x*_ with *x* being the percentage of interactions achieving social distancing, and *C*_*x*_ with *x* being the percentage of symptomatic individuals being confined. Before mitigation, the parameters were {*M*_*18*_, *SD*_*10*_, *C*_*90*_}, on 17 March these changed to {*M*_*14*_, *SD*_*50*_, *C*_*90*_} and on 23-24 March they changed again to {*M*_*7*_, *SD*_*60*_, *C*_*100*_}. The term ‘tuning the model’ means adjusting one or more parameters on one or more days after the lockdown to bring the simulated daily deaths closer to the actual daily deaths. Five contagious individuals were introduced to start an epidemic on day 1. The first mitigation of 17 March was placed when the susceptible population fell to 91%.

### Two modified SIR models

UK Covid-19 mortality data was simulated using a modified classical *SIR* model [10]. For historical interest it is noted that Kermack and McKendrick used *x, y, z* with *K* and *l* where *S,I, R* with *β* and *γ* are now more widely used. The rate of change in the susceptible proportion of the population S, was 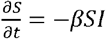 and *β*was modified to represent the mitigations and any weakening, whilst *I* represented the proportion of the population currently infected. Classically 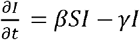 but in this model fell following removal after individuals completed illness periods either after 12.8 days [A] or following two randomly assigned probability distributions [B]. The two distributions were those used in the stochastic model above. Each individual was followed twice, once with a mixture of periods with a random 5% having a longer contagious period to build the epidemic curve, and the other with a random 80% having the longer period to build the mortality curve.

To achieve this, each day had ten equal intervals, Δ*t* = 0.1 *days*. With *n, R(t)* ϵ ℕ, and *S*(*t*),*I*(*t*), *βt* ϵ ℝ we start with *S* (0) = 1, *I*(0) = 1/*n* at *t*= 0. The algorithm proceeds as *t → t +Δt* : S(*t* + Δ*t*) = *S*(*t*) − *βSI* and I(*t +*Δ*t*) = *I*(*t*) + *βSI − R*(*t*)/*n*. For Model 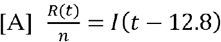. For model [B], two random numbers *µ* and *v*, each with uniform distribution over the interval 0 to 1 were used to look up a time to recovery Φ (µ) from a cumulative probability density function (CDF) for a lognormal distribution. If 0.05 the CDF had meanlog 2.639 and sdlog 0.2, and if *v <* 0.05 the CDF had meanlog 2.99 and sdlog 0.223. In like fashion for both [A] and [B], two random numbers µ and *v*, were used to look up a time to death Ψ (*µ*) from a CDF for a lognormal distribution. If *v ≥* 0.8 the CDF had a meanlog 2.639 and sdlog 0.2, and if *v* < 0.8 the CDF had meanlog 2.99 and sdlog 0.223.

For model [B] we let *k*(*t*) represent the number of new infections, and *R*(*t*) the number of individuals being removed at time We let *d*(*t*) be the number of people dying at time *t* if everyone infected dies. Using [x] to mean the nearest integer less than we have *k*(*t* + *Δt*) = ⌊*nI*(*t* + *Δt*) ⌋, then we increment *R*(*t* + *Δt* + Φ) by 1*k* times with the times, Φ and Ψ, being assessed for each *k* as described. In like fashion we increment *d*(*t* + *Δt* + Ψ) by 1*k* times which is the mortality curve. The shape, but not the magnitude, of this mortality curve was then compared with the shape of the actual daily deaths. In this way a population of twins was followed in which one twin becomes immune and the other dies.

## 3. Results

### The weakening of the lockdown

From around 20 April 2020, actual UK daily deaths gradually rose above the range for simulated daily deaths in the stochastic model [2], (Figure 2, upper panel with arrow). To account for this, it was hypothesised that the lockdown had not been maintained as well as it was when it was first introduced with weakening starting around 7 April, (Figure 2, lower panel with arrow).

**Figure 2.**
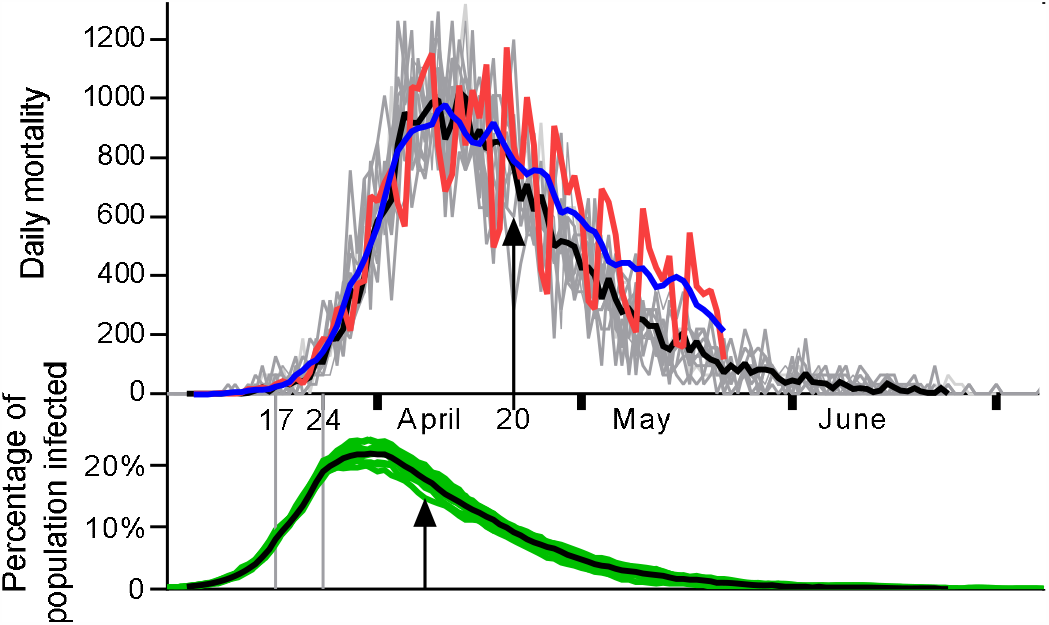
Ten simulations are shown. Actual daily mortality (red, mean blue) and simulated daily deaths (grey, mean black) and percentage of population currently infected (green, with mean black) against time. After 20 April (arrow), mean actual deaths start to exceed mean simulated deaths suggesting a breach of the lockdown, starting perhaps on 7 April (arrow), 13 days previously.

An alternative explanation is that the modelled epidemic curve was too large. This work aimed to (1) tune the stochastic model by making small changes to the parameters defining the lockdown at one or more intervals after 7 April, and (2) model smaller epidemics.

#### Tuning the stochastic model

No single change of any one parameter, on a single date following the lockdown was sufficient to tune the stochastic model. With two changes in the same direction on two separate days tuning was improved. A small number of sets of parameters were then tested on three days. When applied following the lockdown (24 March) on days 16, 23, and 33 respectively, the sets A={*M*_*8*_, *SD*_*60*_, *C*_*90*_}, B={*M*_*9*_, *SD*_*60*_, *C*_*90*_} and C={*M*_*10*_, *SD*_*60*_, *C*_*90*_} produced a promising realignment of simulated and actual daily mortality which was then followed to 27 May, see Figure 3. Only the number of movements per day were increased 7→8→9→10, with all other parameters maintained.

**Figure 3.**
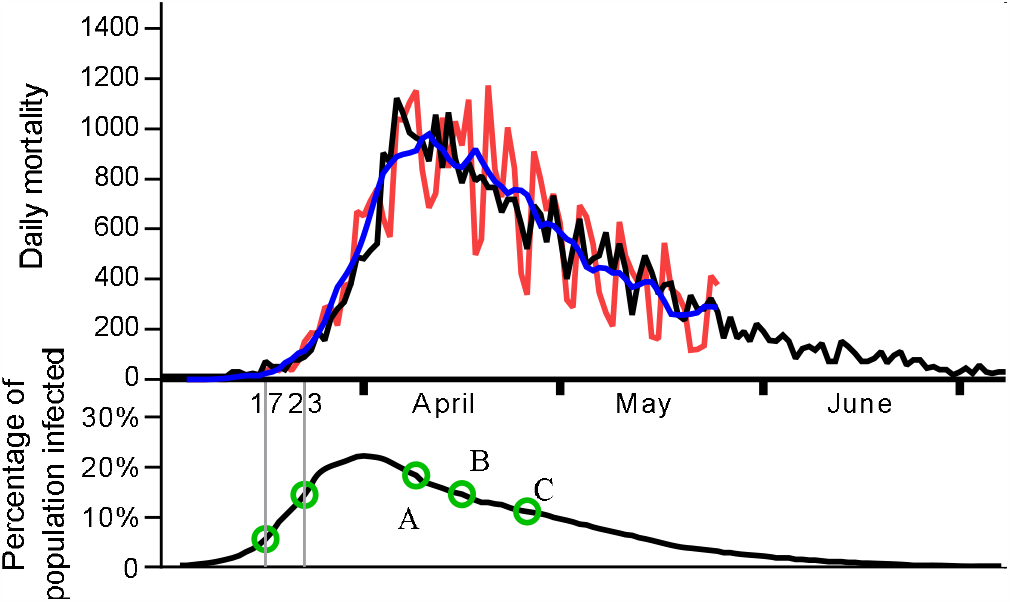
Above: The mean of six simulations with weakening (black) with actual daily deaths (red, smoothed mean blue). Below: The mean mitigated epidemic curve with social distancing 17 March, lockdown 23 March and three step wise weakening changes A, B C.

The parametric changes used to tune the model represent a gradual but progressive weakening of the lockdown in real life. Google mobility data [11] was examined and changes in mobility at the time of the mitigations of 17 and 23–24 March were found. After the lockdown the percentage change from baseline of all relevant activities was similar to the changes required for tuning the model, see Figure 4.

**Figure 4.**
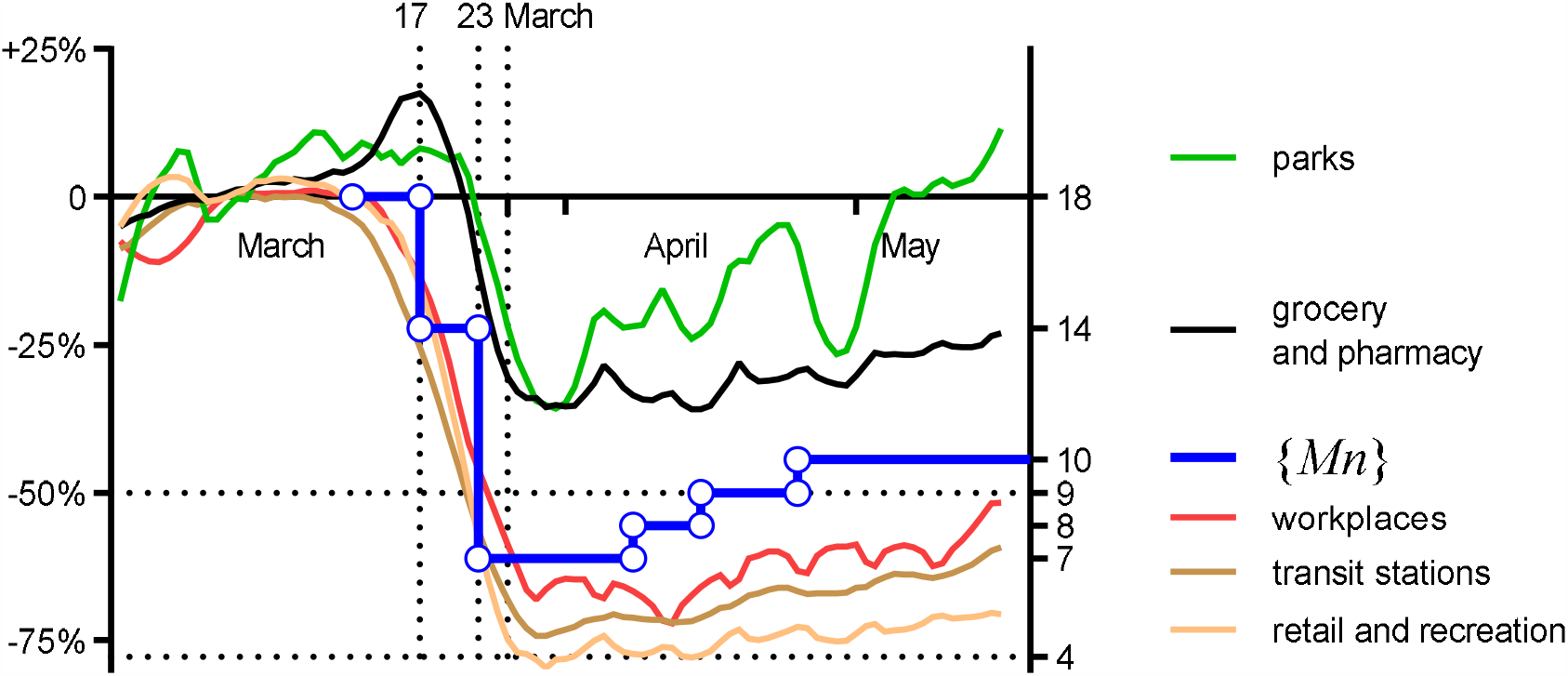
Google mobility data plotted as % change from baseline (left axis) and the movement parameter {M_n_} (blue with circles), which changed in a stepwise fashion (right axis n).

In Figure 4, the percentage change from baseline for UK mobility activity, for 5 domains [11], is shown for the period of the mitigated epidemic, plotted against the left-hand axis as smoothed data. In the Google mobility data, there is evidence of activity gradually, and progressively rising through a few percentage points. On the right-hand axis, the parameter {*M*_*n*_} is plotted such that a fall from 18 to 9 movements a day equates to a 50% fall in baseline activity on the left-hand axis.

Interestingly, the weakening of the lockdown which was modelled to tune the mortality output to fit the actual daily deaths, is similar to the weakening evident in the Google mobility data.

### Locking down earlier or later

A series of epidemics were simulated in the stochastic model with lockdowns starting on different days between 10 March and 1 April. Social distancing was initiated 7 days before each lockdown and the lockdown was weakened in three steps as described above. The total mortality is plotted against the day of the lockdown (not the introduction of social distancing) and appears in Figure 5.

**Figure 5.**
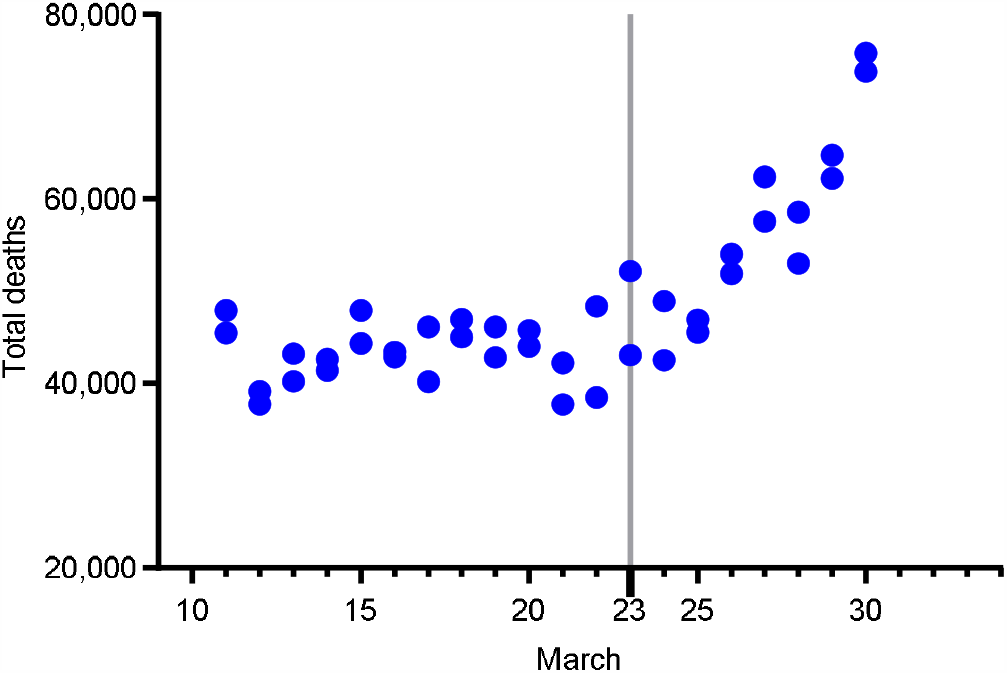
Simulated total deaths from 40 epidemics for lockdowns starting on different days, each with social distancing introduced 1 week earlier and weakening of the lockdown as described. This was output from the stochastic model with social distancing starting with S=91%. The peak in infections was about 20%.

**Figure 6.**
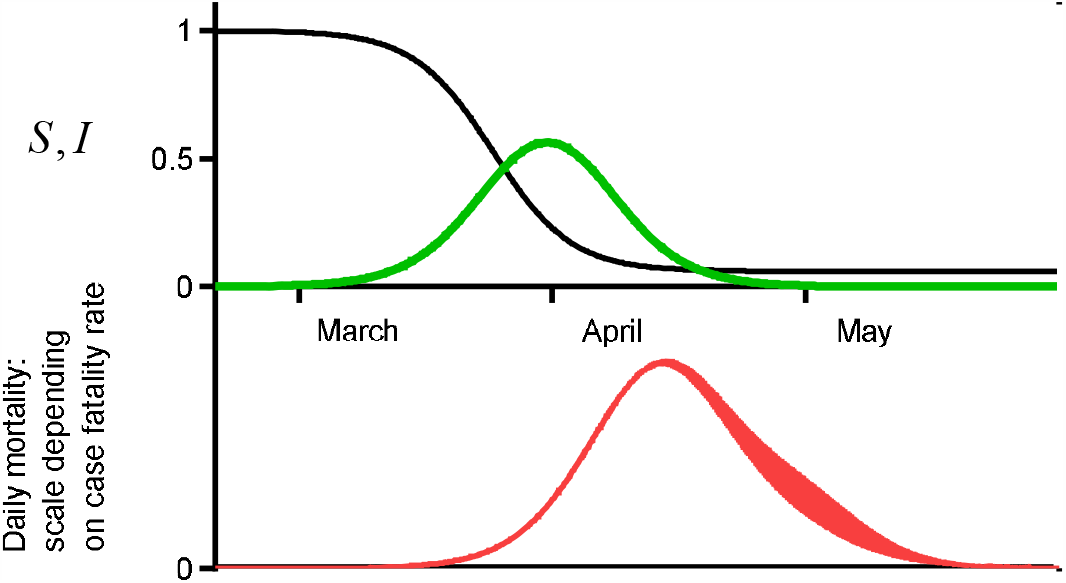
In this unmitigated epidemic 94% of the population became infected: *S* is the susceptible proportion (black), *I* the infected proportion (green) and in red the daily mortality. The area under the curve representing 62,000 deaths with a case fatality rate of 1:1000, and 620,000 with a case fatality rate of 1:100.

In Figure 5 the distribution of data points suggests that an earlier lockdown may not have been associated with fewer deaths, but this is in the context of a significant epidemic curve.

### The two modified SIR models

#### Model [A]

In the modified SIR model [A], a population of 66 million was studied. The value of {3 for an unmitigated epidemic was 0.023 to give a doubling every 3 days, see Error! Reference source not found..

In Model [A], for the doubling to fall appropriately on the introduction of social distancing *β*= 0.01, falling further for the lockdown with *β*= 0.0064. The magnitude of the epidemic curve was altered by changing the start date of the epidemic whilst keeping the two principal mitigations fixed in time. The modelled daily deaths were compared with actual deaths and {3 was altered sequentially with *β*_A_, *β*_B_ and *β* _C_ applied 14, 21 and 28 days after the lockdown to tune mortality.

With a late start and a small epidemic curve, the mortality was a good fit without any weakening of the mitigations. As the epidemic start time was brought earlier and the magnitude of the epidemic curve increased then weakening of the mitigations was required to tune the mortality data.

The largest epidemic in Model [A], with a 19% peak in infections (which does not appear in Figure 7) has its mortality output shown in Figure 8. Earlier lockdowns have significant numbers of deaths.

**Figure 7.**
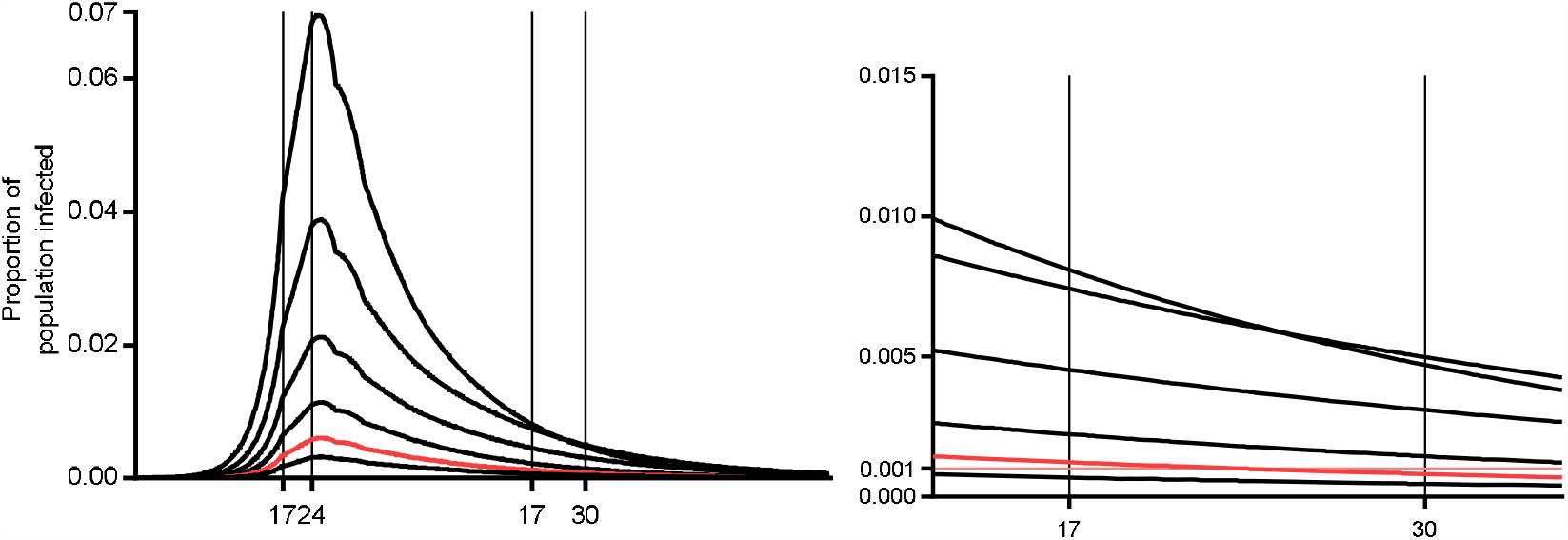
Six epidemic curves for Model [A] showing active cases as a percentage of the population with social distancing 17 March and lockdown 24 March. The percentage infected between 17-30 May appears in the right-hand panel. The ONS data would be consistent with the red epidemic.

**Figure 8.**
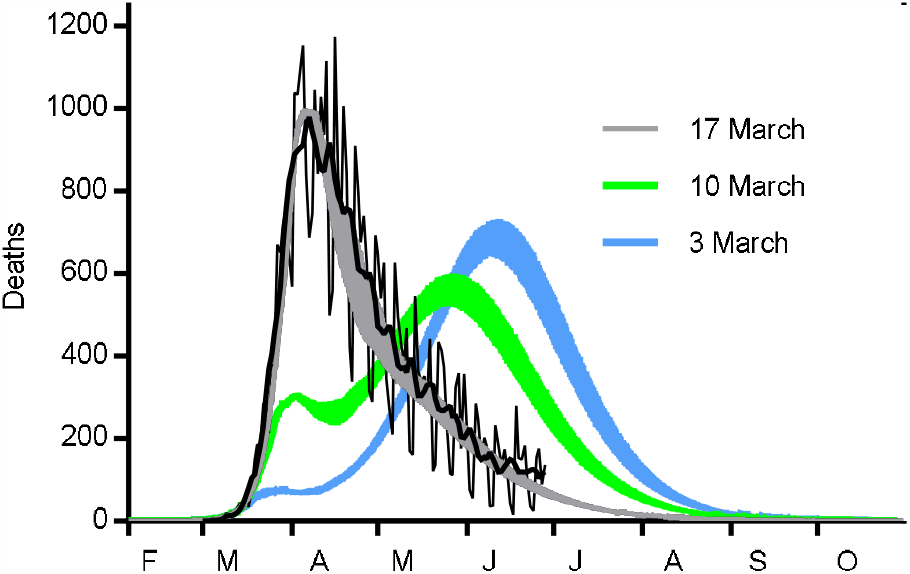
An epidemic in Model [A] starting 2/1/20 with 19% at the peak of infections. Deaths with mitigations starting 17 March (grey), 10 March (green), and 3 March (blue).

The salient features of 10 epidemics with *β*_0_ = 0.023, {*β*3_*sD*_ = 0.01 on the 17 March and *β*_L_ = 0.0064 for the 24 March (except for Epidemic A23 where *β*_*L*_ = 0.0063) appear in Table 2. The designation of an epidemic includes the date in January for the first case: A6 had its first case on 6 January.

**Table 2.** Data from 10 epidemics in Model [A], each initiated with the same *β* changing at the mitigations on 17 and 24 March in the same way. The highlighted cells have the % of active cases found by the Office for National Statistics and Epidemic A14 has a CFR of about 1:100.

| Epidemic [A] | A2 | A5 | A6 | A7 | A8 | A11 | A14 | A17 | A20 | A23 |
| --- | --- | --- | --- | --- | --- | --- | --- | --- | --- | --- |
| $\beta_A$ 7 April | 0.0113 | 0.008 | 0.0076 | 0.0071 | 0.007 | 0.0066 | 0.00645 | 0.0064 | 0.0064 | 0.00635 |
| $\beta_B$ 14 April | 0.0114 | 0.0087 | 0.00815 | 0.0076 | 0.0075 | 0.0068 | 0.00650 | 0.0064 | 0.0064 | 0.00635 |
| $\beta_C$ 21 April | 0.0115 | 0.0095 | 0.0085 | 0.0081 | 0.008 | 0.0071 | 0.00655 | 0.0064 | 0.0064 | 0.00635 |
| Case Fatality Rate | 1:762 | 1:505 | 1:423 | 1:361 | 1:309 | 1:177 | 1:97 | 1:48 | 1:26 | 1:14 |
| Peak Infection | 19% | 12% | 10.0% | 8.4% | 7.0% | 3.9% | 2.1% | 1.1% | 0.61% | 0.32% |
| 17–30 May | 2.93 | 2.23 | 1.62 | 1.4 | 1.34 | 0.72 | 0.38 | 0.22 | 0.13 | 0.07 |
|  | 1.78 | 1.58 | 1.06 | 0.98 | 0.93 | 0.48 | 0.24 | 0.14 | 0.09 | 0.04 |
| 25 May-7 June | 2.17 | 1.81 | 1.25 | 1.1 | 1.07 | 0.56 | 0.29 | 0.16 | 0.10 | 0.05 |
|  | 1.27 | 1.25 | 0.81 | 0.76 | 0.7 | 0.38 | 0.18 | 0.11 | 0.07 | 0.03 |
| S on 1 July 2020 | 49% | 65% | 71% | 75% | 78% | 88% | 93% | 96% | 98% | 98.9% |
| Lockdown 1 week earlier<br>$\beta_{SD}$ 10 March | 47,100<br>(106%) | 43,900<br>(93%) | 31,700<br>(70%) | 28,400<br>(62%) | 25,800<br>(54%) | 15,000<br>(32%) | 12,100<br>(26.3%) | 11,300<br>(24.5%) | 11,100<br>(23.6%) | 10,800<br>(23.1%) |
| Lockdown 2 weeks earlier<br>$\beta_{SD}$ 3 March | 48,900<br>(110%) | 44,500<br>(95%) | 27,800<br>(61%) | 21,700<br>(47%) | 13,200<br>(28%) | 3,900<br>(8.4%) | 2,800<br>(6.2%) | 2,600<br>(5.6%) | 2,500<br>(5.4%) | 2,200<br>(5.2%) |

**Table 3.** Data from 5 Model [B] epidemics, each initiated with the same f3, changing at the mitigation on 17 March to 0.01 but requiring different strengths of lockdown.

| Epidemic [B] | B2 | B5 | B7 | B8 | B11 | B14 |
| --- | --- | --- | --- | --- | --- | --- |
| Lockdown $\beta_L$ from 24 March | 0.0067 | 0.0064 | 0.0064 | 0.0064 | 0.0062 | 0.0058 |
| Weakening $\beta_A$ 7 April | 0.0098 | 0.007 | 0.0066 | 0.00645 | 0.0062 | 0.0058 |
| $\beta_B$ 14 April | 0.0105 | 0.0076 | 0.007 | 0.0066 | 0.0062 | 0.0058 |
| $\beta_C$ 21 April | 0.0115 | 0.0083 | 0.0075 | 0.007 | 0.0062 | 0.0058 |
| Case Fatality Rate | 1:888 | 1:569 | 1:437 | 1:357 | 1:208 | 1:97 |
| Peak in infections | 22.1% | 13.5% | 9.7% | 7.7% | 4.3% | 2.1% |
| 17–30 May | 4.0–2.4 | 2.6–1.7 | 2.3–1.5 | 1.9–1.2 | 1.1–0.72 | 0.5–0.33 |
| 25 May-7 June | 2.9–1.6 | 2.0–1.3 | 1.8–1.2 | 1.5–0.96 | 0.84–0.55 | 0.39–0.26 |
| S on 1 July 2020 | 40% | 65% | 70% | 78% | 86% | 93% |
| Lockdown 1 week earlier $\beta_{SD}$ 10 March | 47,600<br>(106%) | 37,000<br>(81%) | 29,300<br>(63%) | 25,800<br>(54%) | 15,500<br>(33%) | 17,000<br>(37%) |
| Lockdown 2 weeks earlier $\beta_{SD}$ 3 March | 49,200<br>(110%) | 35,000<br>(77%) | 20,700<br>(44%) | 13,200<br>(28%) | 3,900<br>(8.4%) | 2,850<br>(6.2%) |

Epidemics starting earlier had more infections and a lower S when mitigations were introduced. Epidemics starting earlier required greater weakening of the mitigations to tune their mortality curves.

### Model [B]

The salient features for 6 epidemics from Model [B], appear in Table 2: *β*_0_ = 0.023, *β*_*sd*_ = 0.01 on the 17 March with the value for *β* _L_ for the 24 March varying.

Epidemics starting earlier had more infections and a lower S when mitigations were introduced. The mortality curves for 4 Model [B] epidemics appear in Figure 9.

**Figure 9.**
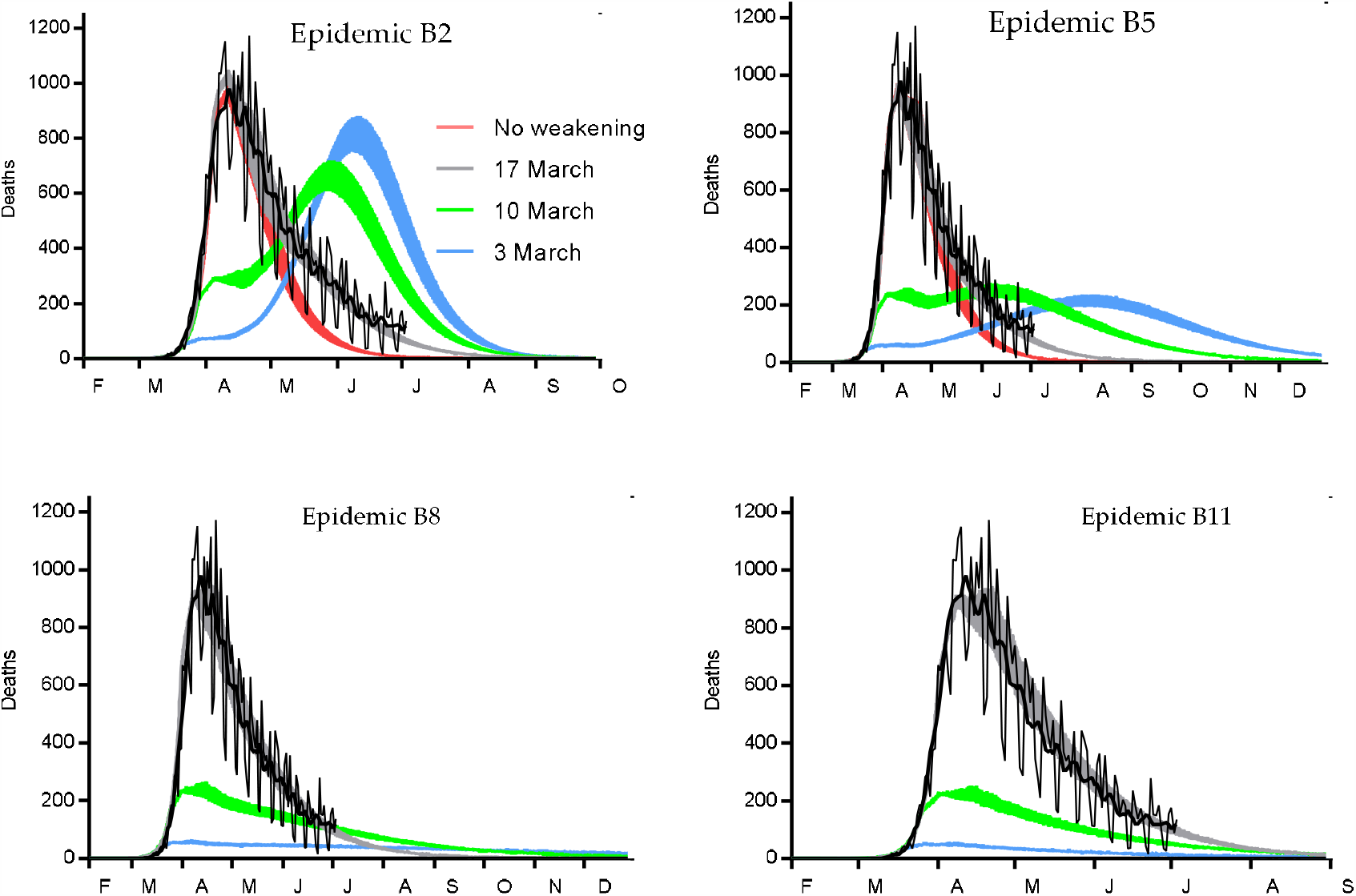
Actual daily UK mortality with smoothed average (black). Four Model [B] epidemics starting on 2–11 January with ever smaller epidemic curves (not shown). Tuned simulated mortality with social distancing 17 March and lockdown 24 March (grey), untuned mortality (red), earlier introduction of social distancing 10 March and lockdown 17 March (green), social distancing 3 March and lockdown 10 March (blue).

There was a linear relationship between the peak % of the population infected and the reciprocal of the case fatality rate; this is shown in two ways in Figure 10.

**Figure 10.**
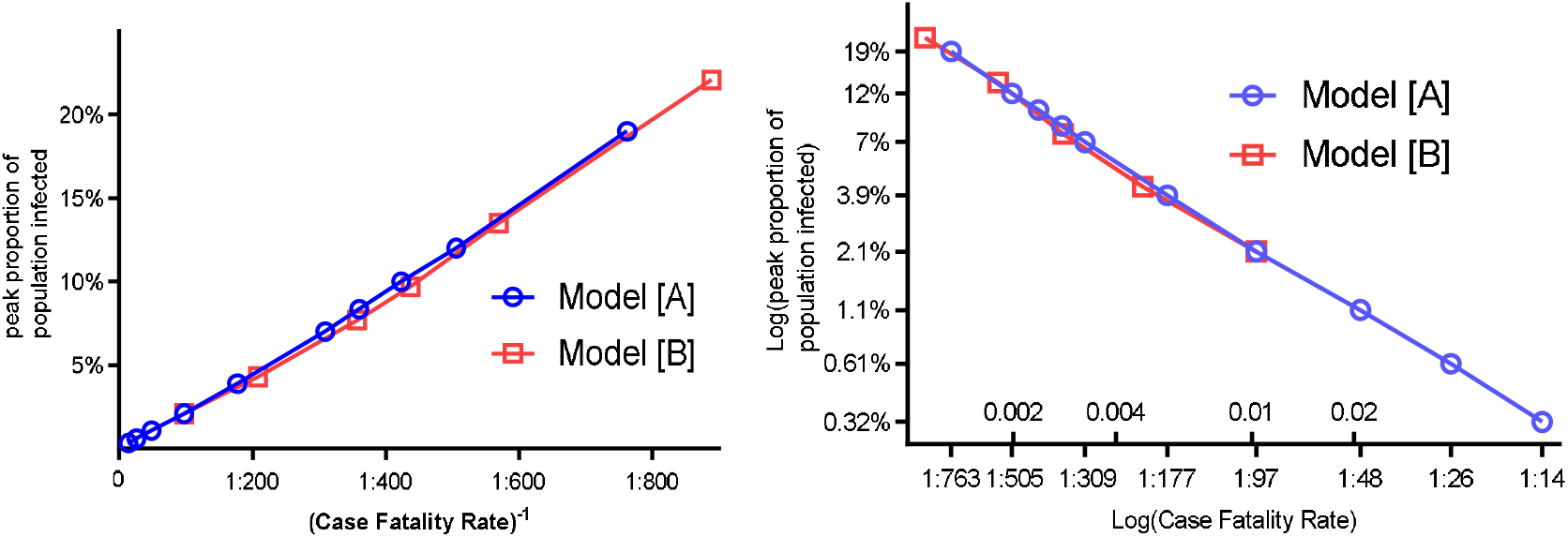
The peak % in *I* is proportional to the reciprocal of the Case Fatality Rate. Model [A] black, Model [B] red.

### Weakening of the mitigations compared with Google mobility data

The parameters required to weaken each of the epidemics were compared with Google mobility data. The comparison is a challenge as we are comparing different phenomena. In Figure 11, Google mobility data is shown with smoothed data as % change from baseline on the left-hand axis for a number of domains. We cannot combine these curves, but together, they give an indication of the depth and timing of changes in activity. On the right-hand axis, with *β* = 0.023 being the base line and the scale logarithmic, the values for weakening of the mitigations are plotted for epidemics A2–A11 in Model [A].

**Figure 11.**
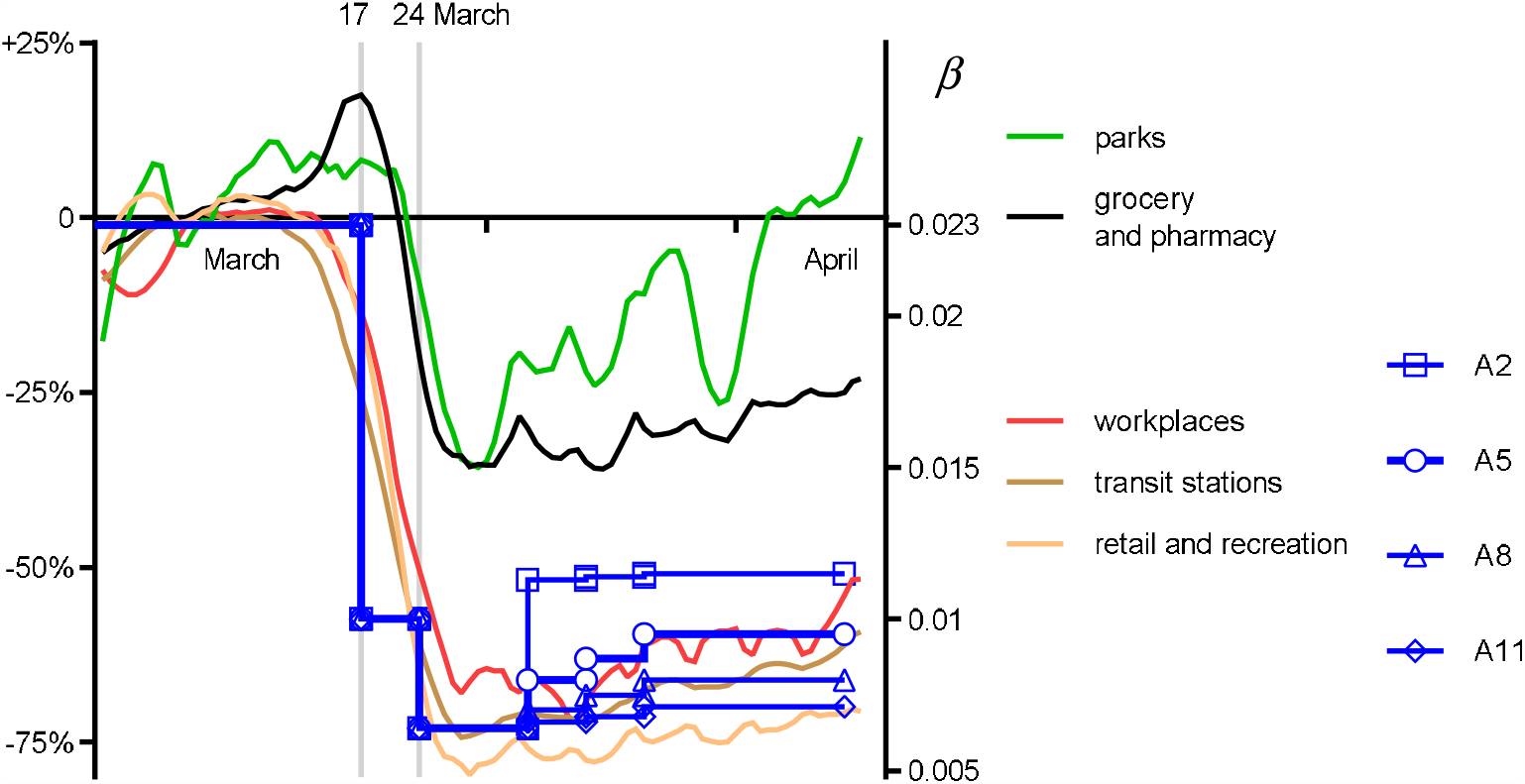
Weakening of the mitigations shown as changes in *β* (left axis) for 4 Model [A] epidemics shown with Google mobility data as % change from baseline (right axis).

In Figure 11 epidemics A5 and A8 are perhaps the most attractive suggesting a CFR between 500 and 300 and peak infections between 12% and 7%. For Model [B] a similar comparison appears in is made in Figure 12.

**Figure 12.**
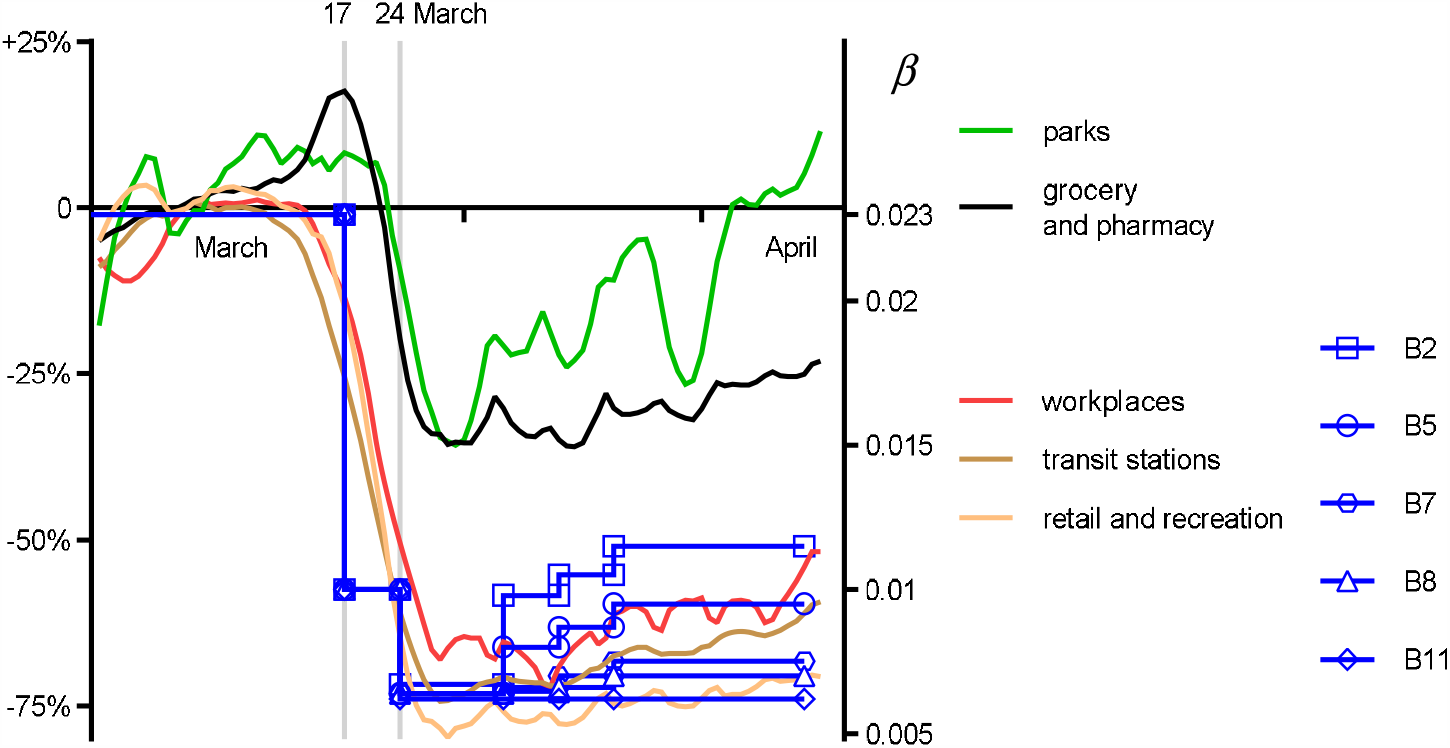
Weakening of the mitigations shown as changes in *β* (left axis) for 5 Model [B] epidemics shown with Google mobility data as % change from baseline (right axis).

In Figure 12, Epidemic B8 and B7 appear to have some consistency with the Google data.

## 4. Discussion and conclusions

A simple stochastic model of the mitigated UK Covid-19 epidemic was developed to simulate daily mortality data and test assumptions about the mechanics of the epidemic and its mitigations. Actual daily deaths started to exceed the mean of simulations and a weakening of the lockdown was hypothesised. With modest but progressive changes in parameters, the model was successfully tuned. The timing and percentage change in the model’s baseline movement parameter was similar to changes published by Google for UK mobility data over the same period.

The principal reason for modelling is to test one’s assumptions about how the real-world works. One may learn more from a failing model than from a model that, by good fortune, produces output that happens to simulate the real world. Models can be criticised if they become over-elaborate or over-parametised or if they are tweaked, simply to produce a desired output. The stochastic model, with its large epidemic curve was tuned to bring its simulated deaths into line with actual deaths by weakening earlier mitigating parameters as the epidemic curve progressed. The impact of the parametric changes on the epidemic curve were necessarily sensitive to the proportion of the population who were susceptible at the time of the change and thereafter. For this reason, advancing or delaying parametric changes had a marked impact on the epidemic curves. Furthermore, it is clear that a small epidemic that is swiftly mitigated, and which barely reduces the susceptible proportion, will be less sensitive to later changes in those parameters compared to larger epidemics which dramatically reduce the susceptible population. If one is to ask about the impact of moving the lockdown on a mitigated epidemic that has weakened, it is crucial to understand the size of the epidemic curve and something about the weakening.

The stochastic model might be considered to have naive parameters making it a poor approximation for human behaviour but the step-wise changes that were introduced to reshape the mortality data have some similarity to real-world changes seen in Google mobility data. If the stochastic model’s parameters are considered naïve then modelling with *β* and particularly *γ* are clearly far more abstract. In the SIR model *β*, has some statistical mechanical elegance in that it relates to a reasonable real-word understanding of the probability of spread of a contagious disease but *γ* is for the same reasons far less attractive. It is for this reason that the fall in *I*, in Model [B], was modelled by Ψ(*µ*).

The modified SIR model that generated an epidemic curve with an average of 0.1% of the population infected between 17-30 May, in line with the ONS figures, was so small that the CFR was 1:26 which seems far too low a figure. The largest modified SIR epidemics had peaks around 20%, close to the stochastic model, and required a significant weakening of the mitigation. For the SIR models this weakening was considered to be more marked than that found in the Google mobility data. Though there is no simple way of determining a relationship between *β* and mobility, except in direction and time, it was felt that epidemics with peak infections between 8–14% and CFR’s 1:300–500 were most likely to share the weakening found in Google’s data.

If the susceptible proportion of the population in early July is as high as this work requires for consistency with the ONS data, or for consistency with a CFR of around 100, then the risks of a second-wave are very high. If the susceptible proportion of the population in early July is lower and consistent with a larger epidemic curve and a lower CFR then the risks of a second-wave are lower.

## Data Availability

All data can be requested from the author.

## Acknowledgments

Thejasvi Subramanian and Louise Lander for contributions to the development of this project.

